# Generation of Synthetic Data in Health Surveys Using Large Language Models

**DOI:** 10.64898/2026.01.27.26345015

**Authors:** David Villarreal-Zegarra, Luciana Bellido-Boza

**Affiliations:** Digital Health Research Center, Instituto Peruano de Orientación Psicológica, Lima, Peru; Universidad Científica del Sur, Lima, Peru; Intendencia de Investigación y Desarrollo, Superintendencia Nacional de Salud, Lima, Peru; Facultad de Ciencias de la Salud, Universidad Peruana de Ciencias Aplicadas, Lima, Peru

**Keywords:** synthetic data, large language models, health surveys, psychometrics

## Abstract

**Background:** Generating synthetic data using artificial intelligence, such as large language models (LLMs), is a useful strategy in public health because it can reduce time and costs, expand access to data, and facilitate information sharing without compromising confidentiality.

**Objective:** To evaluate the consistency and psychometric plausibility of synthetic data generated by an LLM to simulate the responses of survey participants (user personas) in a national health survey in Peru.

**Methods:** We conducted a cross-sectional study based on the National Health Satisfaction Survey (ENSUSALUD 2016) of ambulatory health service users. We used the GPT-OSS-20B model to generate synthetic responses in Spanish, conditioned on narrative profiles derived from sociodemographic and clinical variables. We evaluated consistency between responses and profile characteristics (sex, age, and comorbidities) using performance metrics (accuracy, precision, recall, F1 score, and AUC). We compared distributions between real and synthetic data using t-tests and chi-square tests. For latent variables, we conducted confirmatory factor analyses of the PHQ-9, PHQ-8, and GAD-7 (WLSMV; polychoric matrices) and estimated internal consistency (α and ω). We examined normality (Jarque–Bera test) and stability through correlations between real measures (PHQ-2 and EQ-5D) and synthetic measures (PHQ-2, PHQ-8, PHQ-9, GAD-2, and GAD-7).

**Results:** The model showed strong concordance with the profile for sex, age, and chronic disease status, with metrics close to 1 for most variables; overall consistency was high in the vast majority of cases. The synthetic PHQ-9, PHQ-8, and GAD-7 instruments showed optimal factor fit and high internal consistency. Synthetic measures were positively and significantly correlated with the real PHQ-2 and negatively correlated with EQ-5D, with moderate to high correlations, particularly for PHQ-8/PHQ-9 and GAD-7.

**Conclusions:** An LLM can generate plausible synthetic data for health surveys when its output is conditioned on user personas, preserving high coherence with demographic and clinical characteristics and maintaining adequate psychometric properties in depression and anxiety scales. However, relevant deviations were identified (e.g., overestimation of obesity, unexpected distributions in some variables, and missing values in a sensitive item), which supports the need for rigorous validation and bias control before using these data for inferential purposes or public policy.

## Introduction

The generation of synthetic data using artificial intelligence (AI) is a valuable tool in public health and global biomedical informatics, as it reduces the time and costs required to obtain information, offers high accuracy, and enables researchers to access a wide variety of health-related data [1]. Currently, several AI algorithms enable the generation of synthetic data, including variational autoencoders (VAEs), generative adversarial networks (GANS), and large language models (LLMs). These algorithms have been applied to generate medical images, clinical texts, tabular electronic health record data, and survey responses [2]. In particular, LLMs enable the generation of data that simulate human responses in a coherent, contextualized, and adjustable manner, which is especially useful in health-related contexts [3].

LLMs have been used in health research to create synthetic clinical texts and to simulate patients with specific medical conditions, such as autism or depression [4–6]. For example, ChatGPT has been used to generate clinical narratives of individuals with autism, achieving correct identification by human clinical evaluators in 83% of synthetic cases [6]. Similarly, LLMs have been shown to generate anonymized clinical texts suitable for training clinical named entity recognition (NER) models, even in low-resource languages such as Estonian [5]. In addition, the use of LLMs has been evaluated for generating synthetic data in patient-centered surveys, yielding adequate performance metrics, even with models such as GPT-3.5 [7]. Regarding synthetic survey data generation, a study in the financial domain found that LLM-generated synthetic data achieved 92.6% accuracy in simulated user responses [8]. This demonstrates the effectiveness of LLMs as reproducible, low-cost tools for analyzing hypothetical scenarios. Moreover, LLMs have shown effectiveness in adaptive questionnaires, which condition the model on previous responses to prepare it to present subsequent questions, thereby significantly reducing prediction error and increasing accuracy in question recommendation [9].

The use of LLMs to generate synthetic data in national surveys is a powerful tool; however, it also entails important challenges. Potential challenges include reliance on the reference population, overestimation of reliability, model-specific biases, and possible ethical concerns about sharing generated data with third parties [10]. Furthermore, because different studies employ algorithms optimized for specific tasks, systematic comparisons across studies are difficult. Therefore, independently examining the quality, diversity, and complexity of synthetic data generated by different LLMs is fundamental to avoid underestimating or overestimating the outcomes of interest [11].

An alternative approach to mitigate bias in synthetic data generation is the use of user profiles or LLM-based doppelgänger models, that is, models that incorporate information characterizing participants (sociodemographic variables) and generate responses based on these data [12]. In this context, the present study aims to evaluate the consistency of LLM-based synthetic data for simulating national survey responses using a reference population representative of Peru.

## Method

### Design

This study used a cross-sectional design. It was conducted using data from the National Satisfaction Survey (ENSUSALUD 2016), specifically Questionnaire 1 addressed to outpatient users of the Peruvian health system. This is a secondary database with open access, available through the National Superintendency of Health of Peru (SUSALUD) (http://portal.susalud.gob.pe/blog/base-de-datos-2016).

### Setting and reference population

Peru is a low- and middle-income country with a health system characterized by high fragmentation. The system comprises several subsystems, including the Ministry of Health, the Social Health Insurance (EsSalud), the health services of the armed forces and police, and the private sector. ENSUSALUD 2016 was conducted by the National Institute of Statistics and Informatics (INEI) in collaboration with the National Superintendency of Health (SUSALUD) in 185 health facilities distributed across the 25 regions of the country.

ENSUSALUD 2016 included all adult participants with complete data in the evaluated measurement instruments. Observations with inconsistent data, such as ages greater than 100 years or records with missing information, were excluded. The sample was selected using a probabilistic, nationally representative, two-stage sampling design stratified by geographic region.

### Information on the AI model

We evaluated an open-access large language model, GPT-OSS-20B, developed by OpenAI. This model contains 20 billion parameters and was trained using a diverse corpus of publicly available internet texts. It was released on August 5, 2025. The model corresponds to the base version, without fine-tuning processes or integration with Retrieval-Augmented Generation techniques. It is designed exclusively for text-based tasks and does not support multimodal inputs such as images, audio, or video.

The available hardware consisted of an Intel Core i7-12650H processor, 32 GB of RAM, and an NVIDIA GeForce RTX 3050 GPU. The LLM was executed on a local server using the Ollama architecture. Access to the model was established through a local HTTP endpoint, with standardized execution parameters (temperature 0.0 and a token limit of 2048). Interactions with the model were structured at two levels: a system message defining the user persona or role that the AI model should simulate based on information from ENSUSALUD 2016 users, and a user message containing the questions and response options.

The experimental design allowed evaluation of the LLM’s performance under controlled conditions, recording both accuracy and response consistency. When the LLM did not generate a valid output, either due to deviation from categorical options or responses outside the expected numerical range, an automatic retry was applied using a corrected prompt. If the invalid output persisted, the response was classified as invalid (missing).

### Procedure

#### Creation of a user persona

The process of creating the user persona followed five sequential steps derived from Questionnaire 1 of ENSUSALUD 2016. First, the databases were downloaded, and key variables were identified, integrating sociodemographic dimensions, service utilization, and user perceptions of care. Subsequently, these variables were systematized and relabeled in STATA®, grouped into categories and ranges consistent with the original distribution. Based on this information, narrative profiles in Spanish were constructed to represent realistic archetypes of outpatient users and medical and nursing staff, ensuring diversity and readability. Finally, the user persona and their weighting factors were exported as CSV files.

#### Prompt and creation of synthetic data

The user persona and the questions to be simulated were incorporated using a Python script that automated the combination of each profile with the list of questions defined in CSV files. In each iteration, the model received a system message instructing (prompting) it to “simulate being” the user persona, along with a user message containing the question and response conditions. The system distinguished between categorical and numerical questions. For categorical questions, the output was required to match exactly one predefined option, with automatic validation and retries in case of error. For numerical questions, valid ranges were identified within the question text, and the model was instructed to provide an integer numerical response within those limits. If, after a second attempt, the response was not valid, it was classified as missing to ensure quality control of the generated synthetic data.

### Variables to be generated

#### Patient Health Questionnaire (PHQ-9, PHQ-8, PHQ-2)

The PHQ is an instrument designed to assess depressive symptoms over the previous two weeks, according to the diagnostic criteria established in the Diagnostic and Statistical Manual of Mental Disorders, Fourth Edition (DSM-IV), which were retained in the DSM-5. The scale includes four response options (0 = not at all, 1 = several days, 2 = more than half of the days, 3 = nearly every day) [13]. Several versions exist; the PHQ-9 is the most comprehensive, consisting of 9 items with scores ranging from 0 to 27, whereas the PHQ-8 excludes the suicidal ideation item and yields scores from 0 to 24. In Peru, both versions have demonstrated adequate psychometric properties, including structural validity, internal consistency, age- and sex-based invariance, and sensitivity and specificity [14, 15]. The PHQ-2 focuses exclusively on the two core items (anhedonia and depressed mood), with a score range from 0 to 6, and has been validated in the Peruvian context, showing satisfactory internal consistency [15]. It should be noted that ENSUSALUD only provides information for the PHQ-2.

#### Generalized Anxiety Disorder scale (GAD-7, GAD-2)

The GAD scale assesses anxiety symptoms over the previous two weeks based on DSM-IV criteria, using a Likert-type format with response options ranging from 0 (not at all) to 3 (nearly every day) [16]. The GAD-7, composed of 7 items, has a score range of 0 to 21 and has shown excellent psychometric properties in the Peruvian population under a unidimensional model [15, 17], with adequate internal consistency and evidence of invariance by sex and age [17]. The GAD-2, derived from the GAD-7, focuses on the first two items representing the emotional and cognitive dimensions of anxiety according to the DSM-IV. It has demonstrated acceptable levels of internal consistency (ω = 0.80) and a strong correlation with the extended version in the Peruvian context [15]. It should be noted that ENSUSALUD does not provide information on this variable.

#### Consistency variables

We evaluated variables included in the user persona, such as age (18–99 years) and sex (male or female). In addition, dichotomous variables with “Yes” and “No” response options were included to indicate the presence of common conditions, such as arterial hypertension, type 2 diabetes mellitus, obesity, and depression. The presence of chronic diseases (Yes/No) was also evaluated.

#### Numerical and categorical health variables

We evaluated a set of numerical variables that the AI model can generate as synthetic data, that is, information not available in the original ENSUSALUD database, including height (130– 215 cm) and weight (35–180 kg). We also defined categorical variables, including intelligence quotient (55–145 points), number of hours of nocturnal sleep (3–12 h), and scale scores for single items derived from the PHQ-9 (0–27 points) and GAD-7 (0–21 points). In addition, we summed the synthetic items obtained for each user persona to compute total scores for the PHQ-9 (0–27 points), PHQ-8 (0–24 points), and GAD-7 (0–21 points).

### Analysis plan

#### Consistency analysis

We conducted a consistency analysis to verify that the responses respected the characteristics defined in the user persona profile (sex, age, and presence of a chronic disease). For example, a user persona defined as male should generate responses consistent with sex questions. We analyzed the proportion of cases with responses consistent with the user persona profile. Metrics such as accuracy, precision, recall, F1 score, and AUC were computed.

In addition, we performed a sensitivity analysis to determine whether the distribution of values between synthetic and real data was equivalent. We expected no difference or association between groups. Numerical variables were analyzed using Student’s t-test, whereas dichotomous variables were analyzed using the chi-square test.

#### Psychometric analysis of latent variables

We evaluated coherence in latent variables by examining whether simulated PHQ-9 responses reflected a symptom pattern consistent with the established profile. If the LLM responded appropriately, the outputs should plausibly indicate the presence or absence of depressive symptoms.

We conducted confirmatory factor analysis assuming a unidimensional model for the different versions of the latent scales (i.e., PHQ-9, PHQ-8, and GAD-7). We used the weighted least squares mean and variance adjusted (WLSMV) estimator and polychoric matrices, given their suitability for categorical-ordinal data [20]. Model fit was evaluated using the CFI and TLI (considered adequate if > 0.95), and the SRMR and RMSEA with 90% confidence intervals (adequate if < 0.08) [58]. It was not possible to perform a confirmatory factor analysis for the PHQ-2 and GAD-2, as such analyses require at least 3 items. We expect the synthetic data, as a whole, to show adequate psychometric properties across all cases.

#### Distribution analysis

We analyzed the generated distributions for synthetic numerical variables, such as height and weight, for which an approximately normal distribution is expected. In contrast, for total scores of categorical variables such as the GAD-7 or PHQ-9, an asymmetric distribution is anticipated. Normality was assessed using the Jarque–Bera test. In addition, descriptive analyses were conducted using scatter plots.

#### Stability analysis of synthetic measures

We conducted correlation analyses between PHQ-2 and EQ-5D measurements available for real participants from ENDES 2016 and correlated them with PHQ-2, PHQ-8, PHQ-9, GAD-2, and GAD-7 measurements generated by the AI model. Pearson’s correlation coefficient was used. We expected positive correlations with real PHQ-2 values and negative correlations with EQ-5D. Differences in correlation strength were evaluated using the Williams/Hotelling test.

The EQ-5D is an instrument that measures health-related quality of life and is only available in the ENSUSALUD 2016 dataset. The EQ-5D has demonstrated solid psychometric properties in the Peruvian population [21].

#### Performance metric analysis

Finally, we included operational quantitative metrics such as processing time, tokens used per response, and estimated cost per token. Although the cost depends on the hardware used, electricity consumption, processor efficiency, and other variables, we set the input cost at USD 0.03 per million tokens and the output cost at USD 0.14 per million tokens, based on data from PortKey (https://portkey.ai/models/deepinfra/openai%2Fgpt-oss-20b).

### Ethical considerations

Because this study used a secondary database from the National Superintendency of Health of Peru and did not involve primary data collection, ethics committee approval was not required. Participants in the primary study provided verbal informed consent and voluntarily agreed to participate. Confidentiality and anonymity were ensured, as the database does not contain information that could be used to identify individuals. Minimal risks associated with participation were considered, such as potential boredom or time investment.

## Results

### Consistency analysis

In the consistency analysis, we found that the AI model identified the variables sex, age, and chronic disease status with high concordance, with values of accuracy, precision, recall, F1-score, and AUC close to 1.00 (see Table 1). Similarly, the model showed high performance for depression and type 2 diabetes mellitus, whereas concordance was slightly lower for arterial hypertension (97.3%) and obesity (98.6%), although accuracy and AUC metrics remained high. Notably, high consistency across all cases was observed in 95.9% (n = 927), compared with cases in which at least one variable was inconsistent (4.1%; n = 40). When combining comparisons across all dichotomous variables analyzed for consistency, the LLM demonstrated very high accuracy, precision, F1-score, recall, and AUC values, all close to unity.

**Table 1.** Consistency analysis (n = 1000).

| Variable | Consistent n (%) | Inconsistent n (%) | n | Accuracy | Precision | Recall | F1 | AUC |
| --- | --- | --- | --- | --- | --- | --- | --- | --- |
| Age | 966 (99.9%) | 1 (0.1%) | 967 | 0.99 | - | - | - | - |
| Sex | 1000 (100.0%) | 0 (0.0%) | 1000 | 1.00 | 1.00 | 1.00 | 1.00 | 1.00 |
| Chronic disease | 1000 (100.0%) | 0 (0.0%) | 1000 | 1.00 | 1.00 | 1.00 | 1.00 | 1.00 |
| Depression | 999 (99.9%) | 1 (0.1%) | 1000 | 1.00 | 0.86 | 1.00 | 0.92 | 1.00 |
| Type 2 diabetes mellitus | 1000 (100.0%) | 0 (0.0%) | 1000 | 1.00 | 1.00 | 1.00 | 1.00 | 1.00 |
| Arterial hypertension | 973 (97.3%) | 27 (2.7%) | 1000 | 0.97 | 0.81 | 1.00 | 0.90 | 0.99 |
| Obesity | 986 (98.6%) | 14 (1.4%) | 999 | 0.99 | 0.28 | 1.00 | 0.44 | 0.99 |
| Overall | 5761 (99.3%) | 41 (0.7%) | 5802 | 0.99 | 0.96 | 1.00 | 0.98 | 0.99 |

*Note:* Some values do not sum to 1000 due to missing data. AUC = Area Under the Curve.

The sensitivity analysis did not identify differences in responses across the evaluated groups. That is, incorrectly categorized (inconsistent) data were not associated with synthetic data for dichotomous variables, except for obesity (p = 0.012). In this case, an overestimation of obesity prevalence was observed in the synthetic data group (see Table 2). Upon data review, the 13 additional cases were found to present diabetes (n = 12) or another chronic disease. This suggests that the model may have assumed these conditions were comorbidities of obesity.

**Table 2.** Sensitivity analysis (n = 1000).

|  |  | Real data | Synthetic data |  |
| --- | --- | --- | --- | --- |
| Numerical variables |  | M (SD) | M (SD) | p (t-test) |
| Age |  | 44.61 (16.95) | 44.61 (16.95) | 0.999 |
| Dichotomous variables | Category | n (%) | n (%) | p (chi-square) |
| Sex | Male | 393 (39.3%) | 393 (39.3%) | 1.000 |
|  | Female | 607 (60.7%) | 607 (60.7%) |  |
| Chronic disease | No | 615 (61.5%) | 615 (61.5%) | 1.000 |
|  | Yes | 385 (38.5%) | 385 (38.5%) |  |
| Depression | No | 994 (99.4%) | 993 (99.3%) | 1.000 |
|  | Yes | 6 (0.6%) | 7 (0.7%) |  |
| Type 2 diabetes mellitus | No | 924 (92.4%) | 924 (92.4%) | 1.000 |
|  | Yes | 76 (7.6%) | 76 (7.6%) |  |
| Arterial hypertension | No | 882 (88.2%) | 855 (85.5%) | 0.085 |
|  | Yes | 118 (11.8%) | 145 (14.5%) |  |
| Obesity | No | 994 (99.5%) | 981 (98.2%) | 0.012 |
|  | Yes | 5 (0.5%) | 18 (1.8%) |  |

Note: Some values do not sum to 1000 due to missing data.

### Psychometric analysis of latent variables

Table 3 presents a descriptive summary of the synthetic values generated by the AI model, showing acceptable levels of skewness, kurtosis, and dispersion. In addition, item 9 of the PHQ-9, which assesses suicidal ideation, had the highest number of missing values (n = 301; 30.1%), compared with the remaining items, which ranged from 5 (0.5%) to 19 cases (1.9%).

**Table 3.** Description of PHQ and GAD items (n = 1000).

| Item | 0 = not at all | 1 = several days | 2 = more than half of the days | 3 = nearly every day | M | SD | Skewness | Kurtosis |
| --- | --- | --- | --- | --- | --- | --- | --- | --- |
| **PHQ** |  |  |  |  |  |  |  |  |
| Item 1 | 516 (52.0%) | 123 (12.4%) | 281 (28.3%) | 72 (7.3%) | 0.91 | 1.04 | 0.57 | -1.18 |
| Item 2 | 517 (52.1%) | 167 (16.8%) | 271 (27.3%) | 38 (3.8%) | 0.83 | 0.96 | 0.61 | -1.09 |
| Item 3 | 468 (47.7%) | 112 (11.4%) | 235 (24.0%) | 166 (16.9%) | 1.10 | 1.18 | 0.43 | -1.40 |
| Item 4 | 400 (40.2%) | 107 (10.8%) | 266 (26.7%) | 222 (22.3%) | 1.31 | 1.21 | 0.14 | -1.57 |
| Item 5 | 607 (61.6%) | 203 (20.6%) | 114 (11.6%) | 62 (6.3%) | 0.63 | 0.92 | 1.30 | 0.54 |
| Item 6 | 534 (53.8%) | 159 (16.0%) | 263 (26.5%) | 36 (3.6%) | 0.80 | 0.95 | 0.66 | -1.03 |
| Item 7 | 525 (52.8%) | 113 (11.4%) | 250 (25.1%) | 107 (10.8%) | 0.94 | 1.10 | 0.61 | -1.16 |
| Item 8 | 670 (67.3%) | 109 (11.0%) | 156 (15.7%) | 60 (6.0%) | 0.60 | 0.96 | 1.28 | 0.21 |
| Item 9 | 521 (74.5%) | 137 (19.6%) | 32 (4.6%) | 9 (1.3%) | 0.33 | 0.62 | 2.05 | 4.10 |
| **GAD** |  |  |  |  |  |  |  |  |
| Item 1 | 493 (49.6%) | 331 (33.3%) | 155 (15.6%) | 14 (1.4%) | 0.69 | 0.78 | 0.79 | -0.38 |
| Item 2 | 383 (38.5%) | 179 (18.0%) | 310 (31.2%) | 122 (12.3%) | 1.17 | 1.08 | 0.25 | -1.33 |
| Item 3 | 441 (44.1%) | 261 (26.1%) | 258 (25.8%) | 39 (3.9%) | 0.90 | 0.92 | 0.51 | -0.99 |
| Item 4 | 448 (44.8%) | 189 (18.9%) | 281 (28.1%) | 82 (8.2%) | 1.00 | 1.03 | 0.46 | -1.17 |
| Item 5 | 654 (66.1%) | 227 (22.9%) | 94 (9.5%) | 15 (1.5%) | 0.47 | 0.73 | 1.45 | 1.30 |
| Item 6 | 559 (56.0%) | 115 (11.5%) | 266 (26.6%) | 59 (5.9%) | 0.83 | 1.02 | 0.70 | -1.04 |
| Item 7 | 530 (53.0%) | 306 (30.6%) | 151 (15.1%) | 13 (1.3%) | 0.65 | 0.78 | 0.87 | -0.29 |

Note: Some values do not sum to 1000 due to missing data. PHQ = Patient Health Questionnaire. GAD = Generalized Anxiety Disorder scale.

The PHQ-9, PHQ-8, and GAD-7 showed optimal model fit indices in all cases. In addition, all instruments exhibited high internal consistency coefficients (see Table 4).

**Table 4.** Internal consistency coefficients and internal structure validity of instruments with generated synthetic data (n = 1000).

| Instrument | Items | n | df | $\chi^2$ | CFI | TLI | SRMR | RMSEA [90% CI] | $\alpha$ | $\omega$ |
| --- | --- | --- | --- | --- | --- | --- | --- | --- | --- | --- |
| PHQ-9 | 9 | 660 | 27 | 44.7 | 1.000 | 0.999 | 0.016 | 0.032 [0.013–0.047] | 0.965 | 0.966 |
| PHQ-8 | 8 | 930 | 20 | 36.6 | 1.000 | 0.999 | 0.012 | 0.030 [0.014–0.045] | 0.964 | 0.965 |
| GAD-7 | 7 | 975 | 14 | 18.9 | 1.000 | 1.000 | 0.012 | 0.019 [0.000–0.039] | 0.931 | 0.931 |

Notes: $\chi^2$ = chi-square; df = degrees of freedom; CFI = comparative fit index; TLI = Tucker–Lewis index; SRMR = standardized root mean square residual; RMSEA = root mean square error of approximation. Some values do not sum to 1000 due to missing data.

### Distribution analysis

We identified that only the variable weight, expressed in kilograms, followed a parametric distribution. However, the numerical variable height, which was not part of the user persona information and for which a normal distribution was expected, did not exhibit a parametric distribution (p < 0.05).

In contrast, the remaining variables showed non-parametric distributions (see Table 5). This pattern was expected when using the summed scores of all items from the PHQ-9, PHQ-8, and GAD-7 scales, which exhibited a left-skewed distribution with a higher concentration of cases. However, for the single-item questions assessing depressive and anxiety symptom scores, the distributions showed implausible patterns, characterized by pronounced central peaks and lower frequencies at the extremes (see Figure 1).

**Table 5.** Distribution analysis (n=1000).

| Variable | n | M | SD | Skewness | Kurtosis | JB | p |
| --- | --- | --- | --- | --- | --- | --- | --- |
| Height (cm) (numeric) | 1000 | 167.50 | 6.73 | 0.02 | -0.42 | 7.7 | 0.022 |
| Weight (kg) (numeric) | 1000 | 69.39 | 7.97 | 0.15 | 0.16 | 4.6 | 0.102 |
| Intelligence quotient (categorical) | 993 | 104.92 | 9.18 | 0.63 | 0.44 | 73.1 | <0.001 |
| Hours of sleep (categorical) | 1000 | 6.63 | 0.70 | -0.53 | 0.73 | 67.9 | <0.001 |
| PHQ-9 total (0-27) - Scale | 660 | 7.35 | 7.48 | 0.35 | -1.59 | 82.7 | <0.001 |
| PHQ-8 total (0-24) - Scale | 930 | 7.20 | 7.11 | 0.29 | -1.63 | 115.6 | <0.001 |
| GAD-7 total (0-21) - Scale | 975 | 5.69 | 5.04 | 0.26 | -1.45 | 96.0 | <0.001 |
| Anxiety symptoms (0-21) - Single item | 1000 | 5.75 | 3.44 | 0.05 | -0.71 | 21.5 | <0.001 |
| Depressive symptoms (0-27) - Single item | 1000 | 6.80 | 4.99 | -0.01 | -1.39 | 80.4 | <0.001 |

*Note:* Some values do not sum to 1000 due to missing data. *JB* = Jarque-Bera test was used to assess distributional properties.

**Figure 1.**
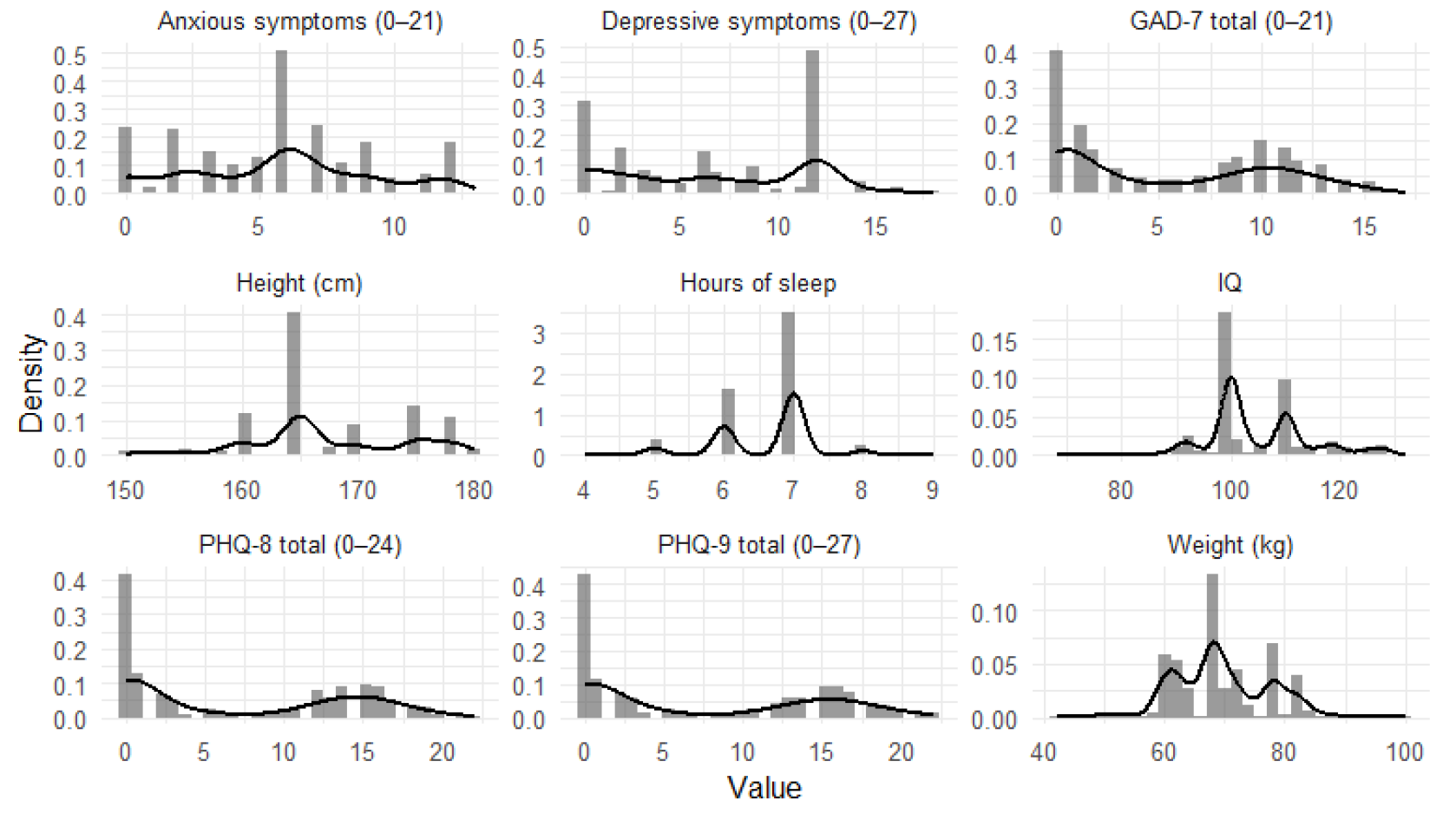
Distribution of the different variables.

### Stability analysis of synthetic measures

We identified significant, positive correlations with moderate to large effect sizes between the different synthetic variables and the real PHQ-2 data. A moderate correlation was observed between real and synthetic PHQ-2 measurements (r = 0.677, 95% CI: 0.642–0.709; n = 987). However, strong, significant, and positive correlations were identified between the real PHQ-2 and the synthetic versions of the PHQ-8 and PHQ-9 (r > 0.80). The remaining correlations are presented in Table 6.

**Table 6.**
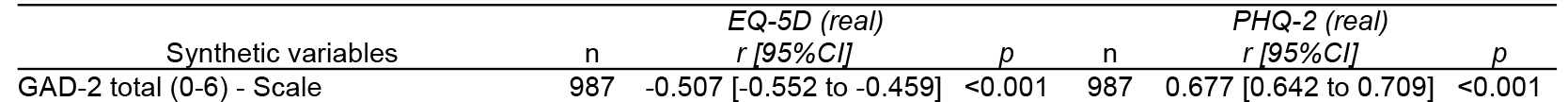
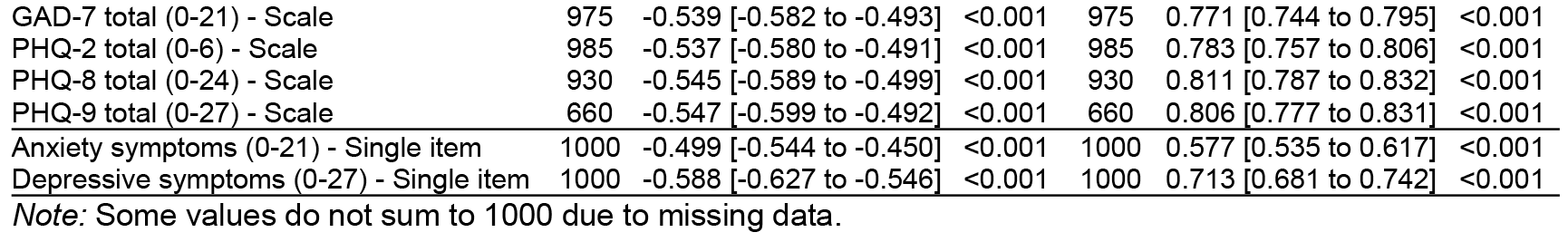
Pearson correlations between real PHQ-2 and EQ-5D values and synthetic data (n=1000).

Moreover, the correlation between the real PHQ-2 and the synthetic PHQ-9 (r = 0.806; 95% CI: 0.777–0.831) was higher than that observed between the real PHQ-2 and the synthetically generated single-item depressive symptom measure (r = 0.713; 95% CI: 0.681–0.742), with a statistically significant difference between correlations of 0.194 (p < 0.001). Similarly, the correlation between the real PHQ-2 and the synthetic GAD-7 (r = 0.771; 95% CI: 0.744–0.795) exceeded that observed with the synthetic single-item depressive symptom measure (r = 0.577; 95% CI: 0.535–0.617), with a statistically significant difference of 0.097 (p < 0.001).

### Performance metric analysis

The mean processing time per question was 5.24 seconds, with an average of 223.66 input tokens and one output token. The mean cost per question (input + output) was USD 0.00000685. Thus, among the 13,814 participants in ENSUSALUD 2016, the total estimated time required to answer a single question was 20 hours, 6 minutes, and 25.36 seconds, with an estimated cost of USD 0.0946231372 per question.

## Discussion

Our study demonstrates that generating synthetic data from user personas in health surveys is feasible and consistent. We successfully synthesized responses for validated instruments (PHQ-8, PHQ-9, and GAD-7) that were not initially available in ENSUSALUD 2016, obtaining robust psychometric properties. In particular, the synthetic data reproduced a clear unidimensional structure for both scales. They showed high internal reliability, with Cronbach’s α coefficients comparable to those reported in real-world applications of these questionnaires [22–25]. This finding is consistent with previous evidence indicating that LLMs can generate coherent medical information that supports imputation processes in health data [26]. For example, recent studies have shown that commercial models (i.e., GPT, Gemini, Cloud) and open-access LLMs can generate realistic synthetic data in health-related contexts, including height, weight, and the prevalence of health conditions, that closely resemble accurate clinical data [27, 28].

In addition, our AI model accurately extracted demographic and clinical information from each profile (sex, age, chronic conditions) and generated responses highly consistent with these characteristics, with accuracy approaching 100% for most simulated variables. This capacity of LLMs to maintain coherence with a given role or profile has been reported by other authors, who note that these models can adopt different “personalities” and respond accordingly at the individual level [29]. Moreover, we observed that synthetic responses correlated strongly with real measurements. For instance, synthetic depression and anxiety scores showed moderate to high positive correlations with real PHQ-2 scores, while exhibiting negative correlations with real EQ-5D quality-of-life measures, as expected. This suggests that the generated data capture plausible clinical patterns, supporting their potential utility. Taken together, our findings indicate that it is possible to obtain high-quality, realistic health survey data from an LLM, thereby expanding its application in public health research.

To our knowledge, no previous studies have simulated national health surveys and evaluated the psychometric properties of synthetic data generated by LLMs to produce representative synthetic estimates. Our results, therefore, address a gap in the literature on the use of synthetic data in population-based surveys. Recent studies have explored partially related applications of LLMs in this field. For example, prior work has shown that LLMs can generate survey responses and mitigate certain wording biases, while emphasizing that they do not resolve sampling or nonresponse issues and should be used in conjunction with traditional methods [29]. Indeed, existing evidence indicates discrepancies between synthetic and real data, as several studies have shown that responses generated by GPT models vary substantially depending on prompt formulation and timing of the query, leading to estimates that differ from those obtained using empirical data [30–32].

A previous study, which did not generate synthetic data but trained a ChatGPT-4 model to administer the PHQ-9 and GAD-7 as an interview, also reported adequate psychometric properties and internal consistency for the LLM’s scores. It should be noted that, whether simulating data or directly evaluating user responses, LLMs are a powerful tool for generating information that appears robust and coherent. However, they remain dependent on prompt design and the specific model employed.

From a global public health perspective, the implications of our findings are substantial. First, the ability to generate realistic synthetic data could transform the survey design and pilot-testing phase. Our results suggest that it is feasible to use an LLM to “respond” to surveys on behalf of simulated participants, allowing researchers to evaluate and refine questionnaires before field implementation, thereby reducing costs and time in preliminary studies. For example, confusing questions or wording biases could be identified in advance by presenting the questionnaire to multiple virtual individuals representing diverse demographic profiles and analyzing their responses. Second, in our study, the model accurately extracted attributes such as age, sex, and health conditions from the profiles and responded consistently based on them. This implies that entire populations could be simulated to anticipate how they might respond to a given survey. This opens the possibility of using synthetic data to optimize item formulation and question sequencing (particularly in adaptive questionnaires) and to test alternative versions of national surveys before involving real participants. Third, generating synthetic data with LLMs offers advantages in terms of equitable access to information and privacy protection. Because this approach does not require identifiable individual-level data, it allows the sharing of datasets resembling real surveys with researchers and policymakers without compromising participant confidentiality. This is particularly relevant in low- and middle-income countries, such as Peru, where access to detailed health data is often restricted due to ethical, logistical, and infrastructural constraints. Synthetic data could be used to fill information gaps, for example, by simulating rare diseases or hard-to-reach subpopulations, enabling hypothesis testing or intervention planning in the absence of sufficient empirical data. Fourth, the inclusion of synthetically generated depression and anxiety scale data could support the training of predictive models or mental health screening systems, even before large-scale real data collection, thereby complementing epidemiological surveillance. Fifth, these applications could accelerate the survey development cycle-from design to validation, and support evidence-informed decision-making using synthetic data when real data are challenging to obtain. Nevertheless, we emphasize that any use in health policy must be accompanied by rigorous quality controls and ethical considerations, as trust in such data depends on demonstrating their validity and the absence of significant biases in each application context.

### Limitations and strengths

This study has several limitations that should be considered when interpreting the results. First, we used only a subset of the information available in ENSUSALUD 2016 (outpatient user and health personnel questionnaires). We did not incorporate the complex survey design (stratification, clustering, or weighting factors) into the generation or analysis of the synthetic data. Although our profiles were based on nationally representative data, the absence of weighting implied an assumption of a uniform distribution of simulated cases.

Second, the generation of synthetic responses relied on a single general-purpose language model (GPT-OSS-20B), without specific fine-tuning for the Peruvian context or health-related terminology. Although the model showed robust overall performance, automatic retries were required when outputs did not precisely match predefined response options or exceeded the allowed numerical ranges. The literature suggests that the quality and fidelity of synthetic data produced by LLMs are highly sensitive to prompt formulation and response constraints. Although we controlled these aspects (e.g., instructing the model to respond only with predefined categories), deviations were still observed in some distributions. For example, the “height” variable did not follow the expected approximately normal distribution, even after conditioning on the profile.

Third, we did not evaluate alternative synthetic data generation methods for comparative purposes, and different approaches may perform differently. This is relevant, as methodological diversity can help avoid both underestimation and overestimation of results. Fourth, although our user persona approach enhances realism by incorporating multiple coherent characteristics per profile, it also introduces dependence on the reference dataset used to construct those profiles. Our system prompts instructed the model to “simulate being” an individual with fixed characteristics (age, sex, medical history), meaning that any bias present in the original ENSUSALUD 2016 data or in the model’s interpretation of these characteristics could be systematically reflected in all responses generated for that profile. In other words, if the model has learned certain biases, these may propagate consistently across the synthetic responses.

Despite these limitations, this study also has notable strengths. To our knowledge, it is the first to simulate responses to a national health survey and conduct a psychometric evaluation using an LLM, demonstrating a novel application of generative AI in a population-based setting. The methodological approach combined the capabilities of an LLM with realistic participant profiles (LLM-based user personas), inspired by recent proposals of “doppelgänger models” that aim to emulate human behavior at the individual level. This allowed us to anchor generated responses in plausible contexts, thereby enhancing their realism. Another strength is the reproducibility and cost-effectiveness of our process, which we achieved through an open-access model and a local execution environment with automated scripts. This means that the methodology can be easily scaled or adapted by other researchers without requiring substantial computational or financial resources.

## Conclusions

In conclusion, our study provides evidence that an LLM can generate plausible synthetic data from health surveys, preserving coherence with the defined characteristics of a reference population and maintaining adequate psychometric properties of clinical scales. These findings open new possibilities for instrument development and validation, as well as for study planning, more efficiently and cost-effectively. However, we emphasize that integrating synthetic data into practice must be approached with caution. Rigorous validation and careful consideration of inherent limitations, such as potential model biases or subtle statistical deviations, are essential before using these data for inference or policy decision-making.

## Data Availability

The generated database, the analysis plan, and other relevant information are available at: https://doi.org/10.6084/m9.figshare.31143748.v1

## Declarations

## Acknowledgments

Not applicable.

## Ethical approval and consent to participate

Because this study used a secondary database from the National Superintendency of Health of Peru and did not involve primary data collection, ethics committee approval was not required. Confidentiality and anonymity were ensured, as the database does not contain information that could be used to identify individuals. This study uses synthetic data from an open-access, anonymized database. Therefore, it does not correspond to a study in humans.

## Consent to publication

Not applicable.

## Declaration of generative AI and AI-assisted technologies in the writing process

We used DeepL to translate specific sections of the manuscript and Grammarly to improve the wording of certain sections. All authors reviewed and approved the final version of the manuscript.

## Authors’ contributions

David Villarreal-Zegarra: Conceptualization, Methodology, Validation, Formal analysis, Investigation, Data Curation, Writing - Original Draft, Visualization.

Luciana Bellido-Boza: Conceptualization, Validation, Investigation, Resources, Writing - Review & Editing, Supervision.

